# Trends and Predictors of Outcomes of Primary Intracerebral Hemorrhage in Very Elderly Patients

**DOI:** 10.1101/2024.06.27.24309617

**Authors:** Kevin Gilotra, Melissa Janssen, Xiaoyue Zhang, Racheed Mani, Sujith Swarna, Cassie Wang, Reza Dashti

**Author notes:** Corresponding Author: Reza Dashti, MD, PhD, Associate Professor, Department of Neurosurgery, Renaissance School of Medicine at Stony Brook University, Cerebrovascular Center, L4-430, Stony Brook, NY 11794-7447, Twitter: @RezaDashtiMD, Facebook: Dr. Reza Dashti. Abbreviations used in this paper: ICH - intracerebral hemorrhage; OR - odds ratio; GCS - Glasgow Coma Scale; HE - hematoma expansion, CI - confidence interval; CT - computerized tomography, DM - diabetes mellitus, HTN – hypertension, HLD – hyperlipidemia, CAD - coronary artery disease, CHF - congestive heart failure, AF - atrial fibrillation, LOS – length of stay, GLMMs - generalized linear mixed effect models, LMMs - linear mixed-effect models.

## Abstract

**Introduction:** Primary intracerebral hemorrhage (ICH) is known to have poor management outcomes. Very elderly patients (age ≥ 80) might have a significantly higher incidence of worse management morbidity and mortality after primary ICH. The aim of this study was to explore presenting status and pre-existing comorbidities in very elderly patients and compare the inpatient management outcomes with younger counterparts.

**Methods:** The Stony Brook ICH database is a retrospective cohort of 814 patients that presented with primary ICH from January 2011 to January 2021. Demographic data, presenting symptoms, pre-existing medical conditions, and imaging findings were recorded. Inpatient outcomes and functional state presented as modified Rankin Scale (MRS) at discharge were evaluated.

**Results:** Our results indicate very elderly patients had significantly higher baseline MRS and comorbidities such as hypertension, hyperlipidemia, and atrial fibrillation at presentation. Similarly, usage of statins, antiplatelets, and anticoagulants were significantly higher in this age group. Very elderly patients were also found to have higher average volume of hematoma at presentation. Our results indicate significantly higher discharge MRS, and inpatient mortality in the very elderly group.

**Conclusion:** Present study demonstrates a wide variety of pre-existing factors that correlate with worse outcomes amongst very elderly patients presenting with primary ICH. Given the importance of aging population as a major healthcare issue in many parts of world, it is crucial to continue exploring these associations in future research. Findings of this study can be utilized to plan further prospective studies on this topic.

## Introduction

Primary intracerebral hemorrhage (ICH) accounts for 10-15% of all strokes and is associated with high management morbidity and mortality rates ^1–4^. While numerous pre-existing comorbidities can put patients at risk for ICH, an increasing age appears to be one of the most remarkable^5^. The global incidence of ICH in general population is predicted as 25 per 100,000 person-years^6^, however, this rate can increase to 136.9 per 100,000 person-years for individuals aged 75-84, and 196.0 per 100,000 person-years for those over the age of 85^6^. Stein et al. proposed a model that predicted a 2.5-fold rise in the proportion of ICH patients over age 80 by the year 2030^1^. As the incidence of ICH increases in the very elderly population, projected healthcare costs are concomitantly rising. Ovbiagele et al. predicted a three-fold increase in the cost of caring for stroke patients over the age of 80 by the year 2030^7^. Patients older than 85 are 10 times more likely to suffer from an ICH than adults aged from 45-54 according to Radholm et al^8^. In addition, adverse outcomes secondary to ICH are more common in the elderly. One study demonstrated a 4-fold increase in 90-day disability and mortality rates when comparing patients older than 75 with those aged over 52^9^. Predictive models currently suggest that ICH-related adverse events will continue to rise in the near future, which is in part due to an aging population, with increasing rates of anticoagulation use and lack of consensus on the best management strategy for elderly^10^.

Given the social and healthcare aspects of population aging, it is of paramount importance to determine the underlying risk factors and predictors of poor outcomes of ICH in elderly patients^5^. Some risk factors for ICH in the general adult population include smoking, cerebral amyloid angiopathy, drug/alcohol use, and pre-existing comorbidities such as hypertension, diabetes mellitus (DM), heart disease and chronic kidney disease^11,12^. In addition, anticoagulation usage is strongly associated with higher mortality and disability rates^10,13^. Most, if not all of these risk factors are likely to be more prevalent in older patients and age itself is one of the greatest independent risk factors for spontaneous ICH^12,14^.

Only a select few studies have evaluated characteristics of very elderly ICH patients and determined risk factors that were associated with poor ICH outcomes while stratifying by age ^1,6,8,15^. However, the results from these studies make it difficult to discern whether the poor outcomes in very elderly patients are due to their age or other pre-existing factors. The current literature lacks consensus on the associations and predictive factors of poor outcomes amongst very elderly ICH patients. The purpose of this study was to compare the characteristics of very elderly ICH patients to younger age groups ICH patients that predict inpatient and discharge outcomes through univariate and multivariate regression analysis.

## Methods

We retrospectively reviewed data from January 2011 to January 2021 for 814 consecutive primary ICH encounters admitted to Stony Brook Hospital with a primary diagnosis of spontaneous ICH. After removing encounters with multiple admissions, 784 unique patients were included in this study. Demographic characteristics, past medical history, therapeutic interventions, and in-hospital complications were assessed for the entire cohort. For our statistical analysis we created two groups: 1. patients over the age of 80 (defined as the very elderly group) and 2. patients aged 18-79. The primary outcomes of interest were inpatient mortality, mRS at discharge, DNR/DNI status, discharge status and hospital length of stay. We categorized all primary outcomes into one of two binary outcomes as either favorable or non-favorable according to the categories described in *Figure 1*. Multiple prior studies have used similar paradigms when classifying favorable and unfavorable outcomes^9,14,16,17^. An mRS score of 0-2 was classified as favorable outcome and mRS of 3-6 as an unfavorable outcome.

Chi-square tests with exact p-values based on Monte Carlo simulation were utilized to compare gender and race/ethnicity distribution between the two groups. Generalized linear mixed effect models (GLMM) considering patients as random effect were used to examine whether there were significant differences in patient characteristics between the two age groups. Similar univariate analyses were conducted to examine the association between variables and binary clinical outcomes. For continuous outcome, i.e. LOS, linear mixed-effect models (LMMs) were conducted, considering patients as random effect. For binary outcomes, multivariable GLMMs were constructed (termed as *Base Models*) which adjusted for variables that had significant associations with their respective primary clinical outcome in the univariate analyses. Separate models were constructed for pre-modified mRS, admission mRS, ICH score, and hematoma volume to fit each outcome if these variables were adjusted in the Base Model. Two-way interaction term between age group and the aforementioned variables were considered in each separate model to examine the possible difference across the levels of the variable in the comparison of the risk of outcome between older and younger groups. For numeric outcome, multivariable LMMs were constructed, following the same process. Participants were treated as the random effect in all GLMMs and LMMs. In GLMMs, an odds ratio (OR) >1 indicated higher risk of outcomes, while an OR <1 indicated lower risk of outcomes. In LMMs, a difference >0 indicated longer LOS, while a difference <0 indicated shorter LOS. Statistical analysis was performed using SAS 9.4 (SAS Institute, Inc., Cary, NC) and the significance level was set at 0.05.

## Results

*Table 1* highlights the demographic characteristics and primary clinical outcomes comparing the young and elderly patients. Of 814 patients admitted for primary spontaneous ICH, 32.4% were over the age of 80 (N=264). We observed an inpatient mortality rate of 25.57% amongst very elderly patients. 10.27% of these patients were discharged home and 93.18% had an mRS at discharge greater than or equal to 3. Hospital length of stay was 3 days shorter in the very elderly group for the univariate analysis (p<0.0001) but demonstrated no significant difference in the multivariate analysis.

*Table 2* highlights patient characteristics for the two groups with p-values to assess significant differences. Medical conditions with a higher prevalence in the elderly group included hypertension (87.1%), hyperlipidemia (60.6%), coronary artery disease (29.8%), atrial fibrillation (32.1%) and cancer (25.9%). Higher rates of medication management, including antiplatelet (59.8%), anticoagulation (29.5%), and statin (46.6%) therapy were observed in the very elderly group compared to the younger group (p-values 0.0006, 0.0246 and 0.0105, respectively). A higher prevalence of obesity was noted in younger patients (38.5%, p-value <0.0001). Although no significant differences were noted in hematoma location between the two groups, very elderly patients were much more likely to present with higher ICH scores (p<0.0001) and higher hematoma volumes (p=0.0220) when compared to the younger group. Additionally, very elderly patients had higher mRS scores at baseline prior to ICH (p=0.0005) and on admission for presentation of ICH (p=0.0438).

*Table 3* demonstrates the base models as a part of the multivariate analysis for each primary outcome (please refer to supplemental tables for details of full model). With regards to primary outcomes, inpatient mortality was not significantly different between the two groups as a part of the univariate analysis. After controlling for mRS on admission, ICH scores, anticoagulation therapy, statin therapy, hematoma volume and history of prior ICH, HLD, CAD, CHF in the multivariate analysis base model, there was again no difference inpatient mortality between the two groups (OR 0.612 95% CI [0.342, 1.095], p=0.0912). A statistically significant difference in the univariate analysis was observed when evaluating mRS at discharge (p=0.0001) and discharge status (p<0.0001) between the two groups. In the multivariate analysis, very elderly patients were more likely to have unfavorable mRS scores at discharge per the base model (OR 0.251 95% CI [0.114, 0.556], p=0.0026) and when adjusting for both mRS scores prior to admission (OR 0.245 95% CI [0.136, 0.445], p<0.001) and at admission. The multivariate analysis for discharge status also demonstrated significant results (OR 0.500 95% CI [0.271, 0.924], p=0.0295) with very elderly patients being less likely to have favorable discharge status after controlling for pre-admission mRS, mRS at admission, ICH scores, hematoma volume, surgical intervention during hospital stay and a history of DM, CAD, AF or statin therapy. Finally, the univariate analysis demonstrated a higher likelihood of DNR/DNI orders placed after admission (p<0.0001) in the very elderly group, although this difference was not present in the multivariate analysis.

## Discussion

Spontaneous ICH remains a devastating disease with poor outcomes that are disproportionately prevalent amongst elderly patients^1,5,18^. The prevalence of spontaneous ICH may increase up to 35.2% by the year 2050, a metric that could be accounted for by both an aging population and increasing rates of anticoagulation for comorbid conditions^1^. Despite this, there is limited literature comprehensively assessing the factors that affect outcomes of ICH in elderly patients. In our study, very elderly patients (age 80 and above) had higher hematoma volumes and ICH scores, more cardiovascular comorbidities, and higher rates of anticoagulation. After performing a univariate analysis that elucidated statistically significant differences in primary outcomes between several patient characteristics, we conducted a multivariate analysis (Tables 3-6) that controlled for these characteristics when comparing very elderly to younger patients. Our multivariate analysis demonstrated that patients over the age of 80 were more likely to have worse mRS scores at discharge and subsequently a higher likelihood of being discharged to rehabilitation centres and acute care facilities. Although very elderly patients demonstrated higher inpatient mortality rates on univariate analysis, the multivariate analysis showed no significant differences in inpatient mortality between the two groups.

Preclinical studies have proposed several pathophysiological mechanisms that may account for poor ICH outcomes in older adults. These include progressive parenchymal degeneration that is more subject to mass effect of hematomas, vascular dysfunction secondary to stiffening, hardening and sclerotic changes in vessels, and the deleterious effects of underlying inflammatory processes due to both non-vascular and cerebrovascular diseases^19^. While the reasons for poor recovery from ICH continue to be investigated in preclinical models, there is a rising need for clinical studies to better understand possible associations that may explain the underlying pathophysiology. In doing so, necessary modifications in the healthcare system can be made to manage the long-term care for elderly ICH patients.

In this study of 814 ICH patients, 32.4% were older than the age of 80. For reference, the INTERACT2 trial, one of the largest landmark studies assessing ICH outcomes in 2794 patients, was comprised of 19.9% of patients over the age of 75^8^. Another large trial of 3448 ICH patients had 34% of patients over the age of 80.^1^. Given that 18.2% of the population in Suffolk County is over the age of 65, this may account for why the proportion of elderly patients in this cohort is much higher in comparison to some other studies^20^. Very elderly ICH patients in this study were also more likely to be of female sex which is in line with Forman et al’s study where 149 of 220 (68%) elderly ICH patients were female^15^. mRS scores both at baseline (p=0.0005) and on admission (p=0.0438) were more likely to be greater than 3 amongst very elderly patients.

The only comorbidity with a higher prevalence in the younger group was obesity, a trend that was also seen in a study by Yang et al.^16^ The remainder of medical conditions which had a significantly higher prevalence in the very elderly group included hypertension (87.1%), hyperlipidemia (60.6%), coronary artery disease (29.8%), atrial fibrillation (32.1%) and cancer (25.9%). The prevalence of these comorbidities in this ICH cohort is significantly higher than both nationwide prevalence rates amongst healthy adults as well as the prevalence rates reported in multiple cohort studies of elderly ICH patients^1,8,16^. This may be accounted for by the fact that our institution primarily serves a suburban, Caucasian population with high rates of access to healthcare insurance and medications needed to manage these comorbidities. As a result, better medical management allows adults to live longer with these comorbidities and may account for why they are more prevalent amongst very elderly ICH patients when compared to their younger counterparts. While medical management of these comorbidities is crucial for improving longevity, one must consider that the presence of cardiovascular disease, malignancy and other comorbidities in the acute setting for an ICH patient is more likely to predict poor outcomes. Therefore, the high complexity of ICH management in elderly patients may in part be due to simultaneously managing their pre-existing and overlapping medical issues, which indirectly contribute to poor outcomes. In our multivariate analysis for each primary outcome, we controlled for the comorbidities noted to have a statistically significant difference for primary outcomes in the univariate analysis.

The higher prevalence of these comorbidities amongst very elderly patients does not necessarily mean they are associated with poor outcomes. Chiquette et al. have also noted multiple characteristics associated with ICH in very elderly patients that upon multivariate analysis, did not seem to predict adverse outcomes^18^. In order to gain a better understanding of the associations seen in both our study and the current literature, longitudinal prospective cohort studies and randomized controlled trials will be needed to further explore associations. Regarding functional outcomes, favorable mRS scores at discharge were extremely rare in the very elderly group as per the multivariate analysis after controlling for relevant covariates. The majority were left markedly impaired with 93.2% having mRS scores greater than or equal to 3 at discharge; a similar metric was seen amongst 558 ICH patients over the age of 75 in the INTERACT2 trial where 74% had an mRS greater than 3^8^. As a result, only 10.3% of very elderly patients in our cohort were discharged home while the rest (89.7%) either died in-hospital or entered acute care facilities or long-term rehabilitation centers due to poor functional outcomes. Similar discharge metric characteristics were reported in Forman et al.’s study where only 11.8% of elderly patients were discharged home and 83.7% either expired or were discharged to hospice, subacute nursing facilities or acute rehabilitation centres^15^. As the prevalence of ICH rises in the aging population, so too will the demand for patient beds in long-term care facilities, as many elderly patients do not have family members at home to care for them long-term^14^. The trends in our study further stress the importance of the future healthcare system allocating more financial resources towards long-term healthcare facilities. In contrast, favorable discharge outcomes rates were significantly higher amongst the younger cohort in this study (p<0.0001) which may be due in part to decreased disability or a higher likelihood of having a family member care for them post-discharge. Notably, the higher prevalence of favorable outcomes was seen even after controlling for important covariates in the multivariate analysis. A relatively favorable prognosis amongst the younger cohort may also explain why these patients had a significantly longer hospital LOS compared to very elderly patients on univariate analysis (p<0.0001).

Patients over the age of 80 were also less likely to undergo neurosurgical intervention and more likely to be made DNR/DNI when compared to younger patients (p<0.0001). Only 7.2% of elderly patients received surgical intervention compared to 21.5% of younger patients. 39% of elderly patients were made DNR/DNI after admission compared to only 22.7% of patients age 18-79. Multiple studies in the literature have suggested that DNR/DNI orders have been associated with worse outcomes due to a lower likelihood of receiving aggressive intervention ^21–25^.

The acute management of ICH currently involves antihypertensives, reversal of anticoagulation and surgical evacuation of the hematoma in case of mass^26^. While variety of surgical procedures for hematoma evacuation and/or external ventricular drain (EVD) placement have all demonstrated efficacy in managing ICH, neurosurgeons might be hesitant to offer surgical intervention to very elderly patients^1,27^. This may be due in part to their poor outcomes despite timely therapeutic measures^28^. With advances in minimally invasive techniques, hematoma evacuation can have increasing role in the treatment of spontaneous ICH^29–32^. While clinical trials are still being conducted, meta-analyses of the current data suggest that these interventions may be associated with lower rates of complications while still offering a mortality benefit. However, the majority of these studies have been conducted in patients under the age of 70, and therefore additional studies will be needed to assess their efficacy in very elderly patients^33^.

Regarding admission laboratory values, a prior study by Inoue et al. demonstrated lower levels of albumin, triglycerides, hemoglobin and alanine aminotransferase in the elderly group amongst a cohort of 377 ICH patients^5^. In our study, we reported no statistically significant differences in routine complete metabolic panel measurements on admission. No differences in complete blood count metrics or coagulation studies were noted on admission either.

Notable strengths of our study include the population size for which ICH was evaluated in older patients. Because ICH is often fatal in older patients, few studies have adequately assessed functional outcomes and long-term mortality in the very elderly. Our series included 264 patients over the age of 80, of which 74.4% were safely discharged. Our results demonstrates that ICH is not always fatal in older populations and that aggressive medical management and surgical interventions may still be used even if the prognosis seems poor. Another important strength of this study is the consideration given to baseline functional status prior to ICH. By utilizing pre-admission mRS, we discerned an effect for functional ability post ICH that took how functional patients were before admission. This is important in an elderly population that is more likely to have worse mRS scores at baseline.

## Limitations

This study has a few limitations. Firstly, the suburban patient population served by Stony Brook University Hospital is more affluent with a greater access to health insurance in comparison to many of the ICH populations studied in the current literature^15^. Our cohort was not very diverse as 87.7% of patients evaluated in this study were of Caucasian race. Therefore, these results may not be easily generalizable to more diverse populations with minority ethnic groups. Additionally, the results also cannot be generalized to populations with ICH who come from lower socioeconomic backgrounds with have limited access to healthcare and poor management of chronic comorbidities. The second limitation of this study is the lack of long-term follow-up. While mRS scores at discharge were obtained, traditional 3-month, 6-month and one year follow-up scores were not obtained as this data was not available for many of these patients who either expired or were lost to follow-up. This makes it difficult to determine long-term prognostic factors for recovery from ICH. Finally, this study is a retrospective analysis performed at an individual hospital which may account for some bias in the results as only one group of patients was analyzed.

## Conclusion

In this study, clinical outcomes at discharge were remarkably worse amongst very elderly patients when compared to their younger counterparts including higher rates of mortality on univariate analysis and more discharges to hospice care and rehabilitation as well as unfavorable discharge mRS scores on multivariate analysis. The results of our study emphasize an increasing need of the healthcare system to facilitate more resources towards long-term care facilities that will inevitably be needed to care for ICH patients with disability^7^. In addition, this study reported a higher prevalence of comorbidities in our patient population in comparison to previous ICH studies which may account for why ICH management in these patients is becoming increasingly complex and costly^7^. Differences in the prevalence of these comorbidities between different ICH populations may have a role in explaining variations in mortality rates and functional outcomes. Longitudinal studies will be needed to further assess the impact of long-term medical management of these outcomes on ICH mortality rates. Altogether, the poor functional outcomes seen in this study, in conjunction with many similar findings in the current literature, further emphasize the need to improve management options for ICH patients over the age of 80.

## Data Availability

“All data referenced in this manuscript is available upon request from the corresponding author.”

## Acknowledgements

We would like to acknowledge all members of the Dashti Lab and Stony Brook University’s Biostatistical Consulting Core who contributed to the research process for this study.

## Appendices

**Figure 1:**
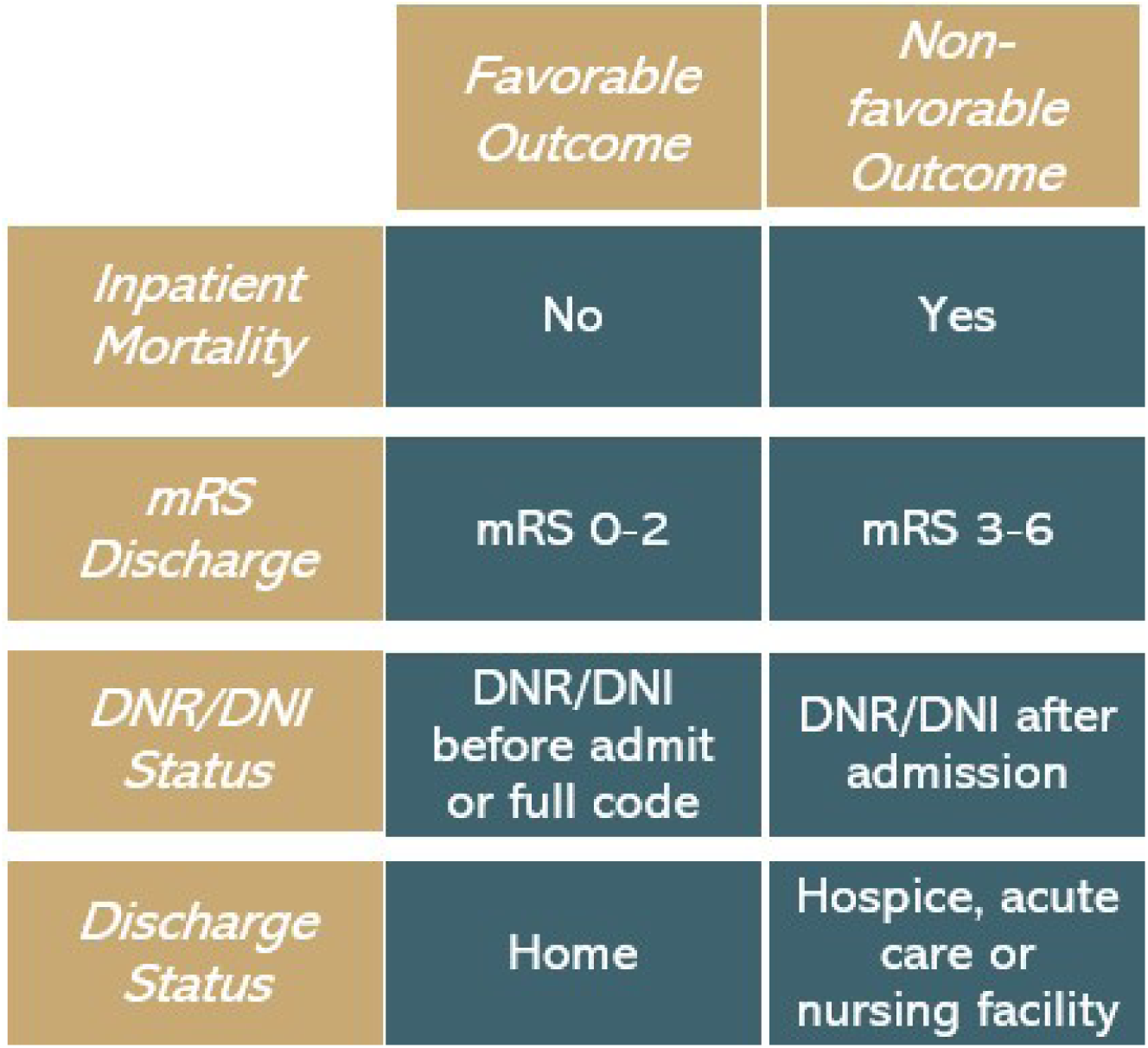
Binary categorization of primary outcomes as either favorable or non-favorable outcomes.

**Table 1:**
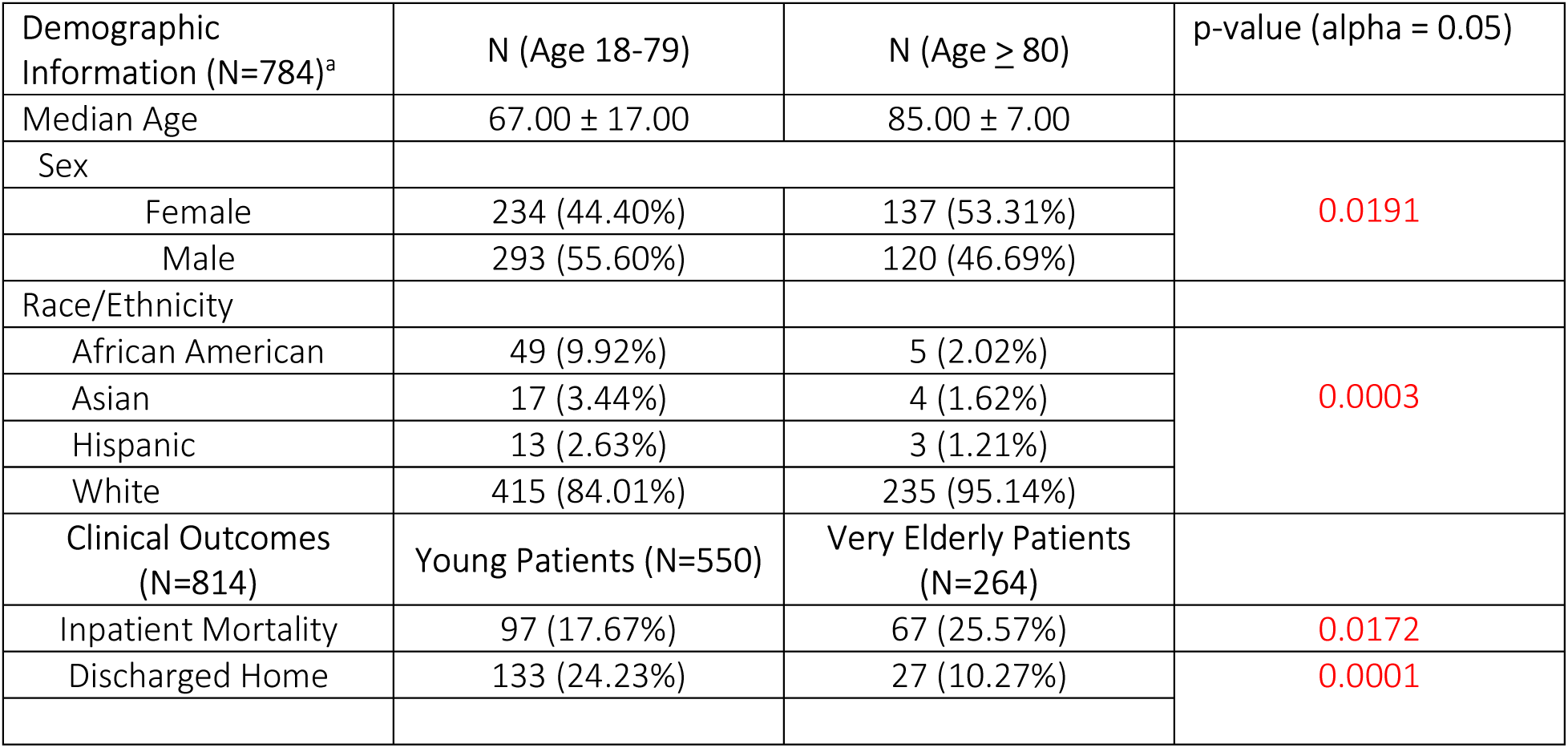
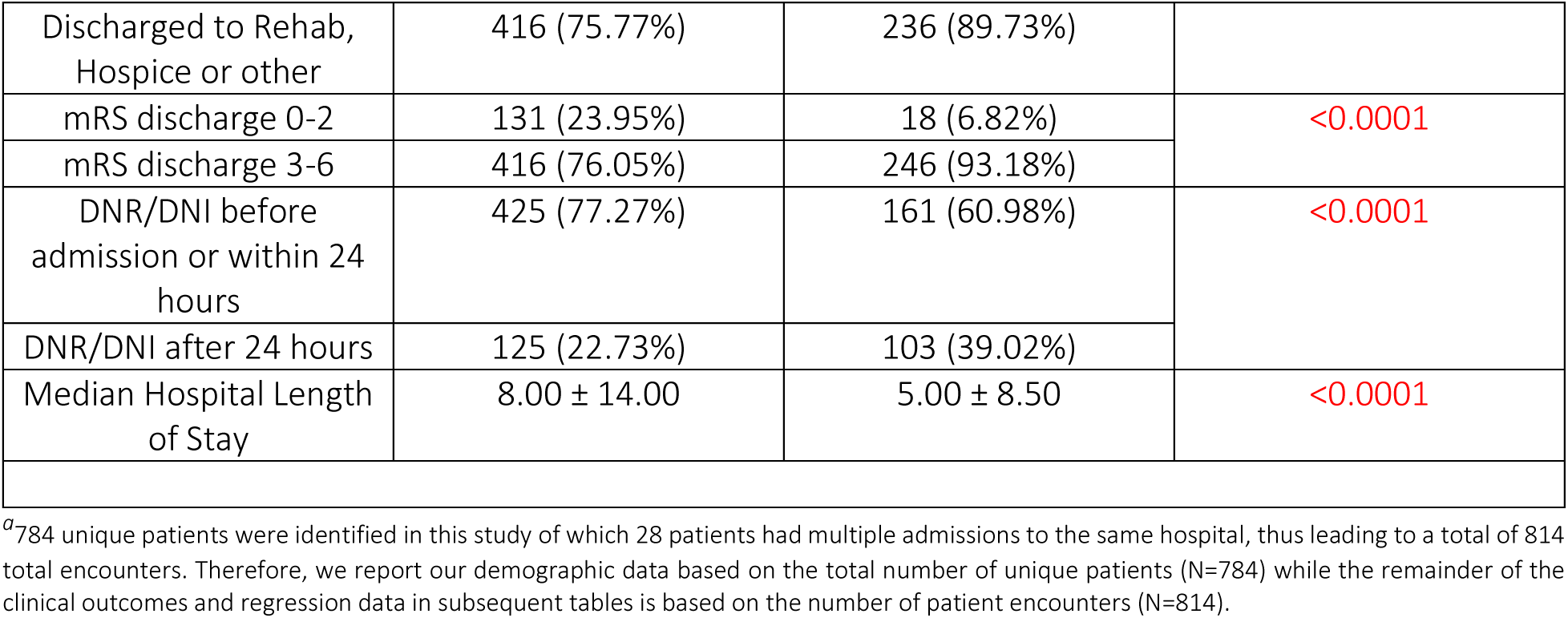
Summary of demographic characteristics and primary outcomes for the very elderly and younger patient groups with p-values based on regression analysis.

| Demographic Information (N=784)<sup>a</sup> | N (Age 18-79) | N (Age ≥ 80) | p-value (alpha = 0.05) |
| --- | --- | --- | --- |
| Median Age | 67.00 ± 17.00 | 85.00 ± 7.00 |  |
| Sex |  |  | 0.0191 |
| Female | 234 (44.40%) | 137 (53.31%) |  |
| Male | 293 (55.60%) | 120 (46.69%) |  |
| Race/Ethnicity |  |  |  |
| African American | 49 (9.92%) | 5 (2.02%) | 0.0003 |
| Asian | 17 (3.44%) | 4 (1.62%) |  |
| Hispanic | 13 (2.63%) | 3 (1.21%) |  |
| White | 415 (84.01%) | 235 (95.14%) |  |
| Clinical Outcomes (N=814) | Young Patients (N=550) | Very Elderly Patients (N=264) |  |
| Inpatient Mortality | 97 (17.67%) | 67 (25.57%) | 0.0172 |
| Discharged Home | 133 (24.23%) | 27 (10.27%) | 0.0001 |
| Discharged to Rehab, Hospice or other | 416 (75.77%) | 236 (89.73%) |  |
| mRS discharge 0-2 | 131 (23.95%) | 18 (6.82%) | <0.0001 |
| mRS discharge 3-6 | 416 (76.05%) | 246 (93.18%) |  |
| DNR/DNI before admission or within 24 hours | 425 (77.27%) | 161 (60.98%) | <0.0001 |
| DNR/DNI after 24 hours | 125 (22.73%) | 103 (39.02%) |  |
| Median Hospital Length of Stay | 8.00 ± 14.00 | 5.00 ± 8.50 | <0.0001 |

<sup>a</sup>784 unique patients were identified in this study of which 28 patients had multiple admissions to the same hospital, thus leading to a total of 814 total encounters. Therefore, we report our demographic data based on the total number of unique patients (N=784) while the remainder of the clinical outcomes and regression data in subsequent tables is based on the number of patient encounters (N=814).

**Table 2:** Characteristics of very elderly group compared to young patient groups including past medical conditions, pre-existing medications, and presenting clinical data based on generalized linear mixed effect models (GLMMs).

| Patient Characteristics | Young Patients (N=550) | Very Elderly Patients (N=264) | p-value |
| --- | --- | --- | --- |
| Hypertension | 148 (18.59%) | 114 (21.39%) | 0.0093 |
| Hyperlipidemia | 361 (45.99%) | 259 (49.24%) | 0.0165 |
| Diabetes | 575 (73.16%) | 378 (71.73%) | 0.2196 |
| Coronary Artery Disease | 612 (77.18%) | 428 (80.60%) | 0.0037 |
| Congestive Heart Failure | 715 (90.16%) | 481 (90.58%) | 0.5855 |
| History of Previous Intracranial Hemorrhage | 703 (88.65%) | 468 (87.97%) | 0.4161 |
| Atrial Fibrillation | 616 (77.78%) | 438 (82.64%) | 0.0001 |
| Cancer | 627 (79.27%) | 433 (81.85%) | 0.0190 |
| BMI | 519 (67.49%) | 324 (61.48%) | <.0001 |
| **Medications** |  |  |  |
| Statin | 491 (60.32%) | 350 (63.64%) | 0.0105 |
| Anticoagulant | 593 (75.93%) | 411 (78.59%) | 0.0246 |
| Antiplatelet | 409 (50.25%) | 303 (55.09%) | 0.0006 |
| **Presenting Clinical Data** |  |  |  |
| *Hematoma Location* |  |  |  |
| Deep Supratentorial | 233 (43.47%) | 97 (37.74%) | 0.0846 |
| Lobar | 266 (49.63%) | 149 (57.98%) |  |
| Other | 37 (6.90%) | 11 (4.28%) |  |
| Median Hematoma Volume | 11.50 ± 34.00 | 14.33 ± 42.95 | 0.0220 |
| *ICH Score* |  |  |  |
| 0 | 172 (31.27%) | 0 (0.00%) | <.0001 |
| 1 | 161 (29.27%) | 83 (31.56%) |  |
| 2 | 114 (20.73%) | 73 (27.76%) |  |
| 3 | 77 (14.00%) | 61 (23.19%) |  |
| 4 | 26 (4.73%) | 29 (11.03%) |  |
| 5 | 0 (0.00%) | 17 (6.46%) |  |
| *Pre-admission mRS* |  |  |  |
| 0-2 | 466 (86.94%) | 191 (75.20%) | 0.0005 |
| 3-6 | 70 (13.06%) | 63 (24.80%) |  |
| *mRS at Admission* |  |  |  |
| 0-2 | 94 (17.54%) | 30 (11.54%) | 0.0438 |
| 3-6 | 442 (82.46%) | 230 (88.46%) |  |

**Table 3:** Odds ratio with 95% CIs for binary outcomes by age group based on GLMMs with multivariate analysis.

| Model | Variable | Level | OR (95% CI) | P-value<sup>1</sup> |
| --- | --- | --- | --- | --- |
| **Outcome: Inpatient death<sup>2</sup>** |  |  |  |  |
| Base model<br>Controlled for: MRS at admission, ICH score, HLD, CAD, CHF, History of Previous ICH, Obesity (BMI>30), statin use, anticoagulant use and hematoma volume | Age | ≥80 vs 18-79 | 0.612 (0.342, 1.095) | 0.0912 |
|  | MRS at admission | 3-6 vs 0-2 | 5.333 (1.372, 20.727) | 0.0195 |
|  | ICH score | 3-6 vs 0-2 | 7.613 (4.233, 13.690) | <.0001 |
|  | Hematoma volume | Unit = 1 mm<sup>3</sup> | 1.003 (0.998, 1.009) | 0.2227 |
|  | Age * ICH score | ≥3 vs <3<br>[Age≥80] | 10.215 (4.519, 23.092) | <.0001 |
| **Outcome: Favorable discharge status<sup>3</sup>** |  |  |  |  |
| Base model<br>Controlled for: pre-modified mRS, mRS at admission, ICH score, surgical intervention, DM, CAD, AF, statin use, and hematoma volume | Age | ≥80 vs <80 | 0.500 (0.271, 0.924) | 0.0295 |
|  | Pre-modified MRS | 3-6 vs 0-2 | 0.761 (0.353, 1.642) | 0.4614 |
|  | MRS at admission | 3-6 vs 0-2 | 0.143 (0.081, 0.251) | <.0001 |
|  | ICH score | 3-6 vs 0-2 | 0.152 (0.048, 0.482) | 0.0034 |
|  | Hematoma volume | Unit = 1 mm<sup>3</sup> | 0.995 (0.986, 1.004) | 0.2546 |
| **Outcome: Favorable MRS at discharge<sup>4</sup>** |  |  |  |  |
| Base model<br>Controlled for: pre-modified mRS, mRS at admission, ICH score, surgical intervention, HTN, DM, CAD, CHF, AF, statin use, anticoagulant use and hematoma volume | Age | ≥80 vs <80 | 0.251 (0.114, 0.556) | 0.0026 |
|  | Pre-modified MRS | 3-6 vs 0-2 | 0.060 (0.006, 0.558) | 0.0177 |
|  | MRS at admission | 3-6 vs 0-2 | 0.082 (0.043, 0.156) | <.0001 |
|  | ICH score | 3-6 vs 0-2 | 0.093 (0.018, 0.477) | 0.0081 |
|  | Hematoma volume | Unit = 1 mm<sup>3</sup> | 0.996 (0.985, 1.006) | 0.3608 |

*Outcomes in very elderly patients with primary ICH*
| Model | Variable | Level | OR (95% CI) | P-value<sup>1</sup> |
| --- | --- | --- | --- | --- |
| **Outcome: Unfavorable time to DNR/DNI<sup>5</sup>** |  |  |  |  |
| Base model<br>Controlled for: pre-modified mRS, mRS at admission, ICH score, hypertension, CAD, atrial fibrillation, BMI, anticoagulant use and hematoma volume | Age | ≥80 vs <80 | 1.251 (0.781, 2.003) | 0.3262 |
|  | Pre-modified MRS | 3-6 vs 0-2 | 1.218 (0.710, 2.089) | 0.4478 |
|  | MRS at admission | 3-6 vs 0-2 | 3.251 (1.381, 7.654) | 0.0102 |
|  | ICH score | 3-6 vs 0-2 | 5.444 (3.304, 8.972) | <.0001 |
|  | Hematoma volume | Unit = 1 mm<sup>3</sup> | 1.000 (0.995, 1.006) | 0.8447 |
| 1: P-values were from generalized linear mixed effect models with patients' ID as random effect.<br>2: Complete model results were listed in supplemental Table 9.<br>3: Complete model results were listed in supplemental Table 10.<br>4: Complete model results were listed in supplemental Table 11.<br>5: Complete model results were listed in supplemental Table 12. |  |  |  |  |

**Table 4:** Mean admission laboratory values in each age group. For categorical variables, column percentages were presented.

| Variable | Level | Age <80 | Age ≥80 |
| --- | --- | --- | --- |
| gfrBL | 0 - 30 | 38 (7.14%) | 9 (3.52%) |
|  | 30 - 60 | 79 (14.85%) | 81 (31.64%) |
|  | 60 - 120 | 405 (76.13%) | 165 (64.45%) |
|  | >120 | 10 (1.88%) | 1 (0.39%) |
| GFRmin | 0 - 30 | 58 (10.90%) | 17 (6.64%) |
|  | 30 - 60 | 133 (25.00%) | 102 (39.84%) |
|  | 60 - 120 | 338 (63.53%) | 137 (53.52%) |
|  | >120 | 3 (0.56%) | 0 (0.00%) |
| Sodium on admission |  | 139.28 ± 4.07 | 139.39 ± 4.32 |
| minimumNa |  | 136.03 ± 4.23 | 136.93 ± 4.87 |
| maximumNa |  | 145.50 ± 7.74 | 143.95 ± 7.05 |
| Chloride on Admission |  | 102.55 ± 5.31 | 102.64 ± 4.65 |
| baseline glucose |  | 147.54 ± 69.96 | 139.30 ± 44.81 |
| Baseline Creatinine |  | 1.12 ± 1.20 | 0.95 ± 0.38 |
| Max Creatinine |  | 2.05 ± 8.96 | 1.12 ± 0.61 |
| Calcium on admission |  | 9.00 ± 0.75 | 9.05 ± 0.60 |
| Phosphorus on admission |  | 3.23 ± 0.88 | 3.10 ± 0.66 |
| Magnesium on admission |  | 1.95 ± 0.31 | 1.92 ± 0.29 |

*Outcomes in very elderly patients with primary ICH*
| Variable | Level | Age <80 | Age ≥80 |
| --- | --- | --- | --- |
| max troponin |  | 0.13 ± 0.75 | 0.11 ± 0.70 |
| HgBA1c |  | 6.03 ± 1.35 | 5.80 ± 0.67 |
| WBC |  | 10.40 ± 4.33 | 9.95 ± 4.22 |
| hemoglobin |  | 13.19 ± 2.08 | 12.87 ± 1.66 |
| platelet count on admission |  | 228.28 ± 79.23 | 215.93 ± 77.74 |
| INR on admission |  | 1.27 ± 0.86 | 1.26 ± 0.77 |
| ptt on admission |  | 30.65 ± 7.35 | 31.14 ± 7.19 |

